# A genome epidemiological study of SARS-CoV-2 introduction into Japan

**DOI:** 10.1101/2020.07.01.20143958

**Authors:** Tsuyoshi Sekizuka, Kentaro Itokawa, Masanori Hashino, Tetsuro Kawano-Sugaya, Rina Tanaka, Koji Yatsu, Asami Ohnishi, Keiko Goto, Hiroyuki Tsukagoshi, Hayato Ehara, Kenji Sadamasu, Masakatsu Taira, Shinichiro Shibata, Ryohei Nomoto, Satoshi Hiroi, Miho Toho, Tomoe Shimada, Tamano Matsui, Tomimasa Sunagawa, Hajime Kamiya, Yuichiro Yahata, Takuya Yamagishi, Motoi Suzuki, Takaji Wakita, Makoto Kuroda, COVID-19 Genomic Surveillance Network in Japan

## Abstract

**Background:** After the first case of COVID-19 in Japan on 15 January 2020, multiple nationwide COVID-19 clusters were identified by the end of February. The Japanese government focused on mitigating emerging COVID-19 clusters by conducting active nationwide epidemiological surveillance. However, an increasing number of cases appeared until early April, many with unclear infection routes exhibiting no recent history of travel outside Japan. We aimed to evaluate the severe acute respiratory syndrome coronavirus 2 (SARS-CoV-2) genome sequences from COVID-19 cases until early April and characterise the genealogical networks to demonstrate possible routes of spread in Japan.

**Methods:** Nasopharyngeal specimens were collected from patients and a quantitative reverse transcription polymerase chain reaction testing for SARS-CoV-2 was performed. Positive RNA samples were subjected whole genome sequencing and a haplotype network analysis was performed.

**Findings:** Some of the primary clusters identified during January and February in Japan directly descended from Wuhan-Hu-1-related isolates in China and other distinct clusters. Clusters were almost contained until mid-March; the haplotype network analysis demonstrated that COVID-19 cases from late March through early April may have caused an additional large cluster related to the outbreak in Europe, leading to additional spread within Japan. National self-restraint during February was effective in mitigating the COVID-19 spread, but late action on stopping immigration and declaring national emergency in Japan might be involved in the later increase in cases.

**Interpretation:** Genome surveillance suggested that at least two distinct SARS-CoV-2 introductions from China and other countries occurred.

**Funding:** Japan Agency for Medical Research and Development.

## Introduction

The initial coronavirus disease 2019 (COVID-19) outbreak occurred in Wuhan, China in late December 2019. It was caused by a new strain of beta coronavirus, known as the severe acute respiratory syndrome coronavirus 2 (SARS-CoV-2) ^1-3^. After the identification of the first patient with COVID-19 in Japan on 15 January 2020, multiple local COVID-19 clusters were identified nationwide by the end of February. The Japanese government focused on identifying and mitigating emerging COVID-19 clusters before they could spread further. In an effort to contain these clusters and limit the number of new cases, active, nationwide epidemiological surveillance of each cluster was conducted in order to identify the close contacts of existing patients with COVID-19. Japan has sustained a moderate spread by focusing on COVID-19 outbreak clusters; however, an ever-increasing number of COVID-19 cases appeared until early April, which made it difficult to identify all infection routes.

Although some of the COVID-19 clusters were successfully contained, the number of cases continued to increase. As of 16 April, the Japanese government declared a nationwide state of emergency in view of the worsening spread. To support the ongoing epidemiological surveillance, we collaborated with the local public health institutes in Japan (Supplementary Table S1) and conducted a whole genome sequencing of SARS-CoV-2. Our goal was to apply genomic epidemiology to predict potential routes of infection within or between clusters ^4^. Thus far, multiple studies have been conducted to demonstrate the COVID-19 spread under nationwide and global comparative genome surveillance; as of 2 June, there were already 33,483 complete genomes publicly available in the Global Initiative on Sharing All Influenza Data (GISAID) platform ^5^. Some detailed reports have been conducted to highlight regional specific clusters and its intra- and international spreads in the US ^6^, UK ^7^, Hungary ^8^, Australia ^9^, Denmark ^10^, Iceland ^11^, and California ^12^

In this study, we aimed to evaluate viral genome sequences from COVID-19 cases until early April and characterise the genealogical networks to demonstrate possible routes of spread in Japan.

## Methods

### Ethical approval and consent to participate

The study protocol was approved by the National Institute of Infectious Diseases in Japan (Approval No. 1091). It was conducted according to the principles of the Declaration of Helsinki, in compliance with the Law Concerning the Prevention of Infections and Medical Care for Patients of Infections of Japan. The ethical committee waived the need for written consent since the study involved the viral genome sequencing. The personal data related to the clinical information were anonymised, and our procedure is to not request written consent for all patients with COVID-19.

### Clinical specimens and reverse transcription polymerase chain reaction (RT-qPCR) testing for COVID-19

Nasopharyngeal specimens were collected from patients and a quantitative RT-qPCR testing for SARS-CoV-2 ^13,14^ was performed at local public health institutes in Japan (Supplementary Table S1). The positive RNA samples were subjected to whole genome sequencing.

### Whole genome sequencing of SARS-CoV-2

Whole genome sequences of SARS-CoV-2 were obtained by means of the PrimalSeq protocol for enriching the cDNA of the SARS-CoV-2 genome using multiplex RT-PCR, as proposed by the Wellcome Trust ARTIC Network ^15^. We found two amplicons that regularly showed low to zero coverage due to primer dimerization as described by Itokawa et al. ^16^. Therefore, we have modified the ARTIC Network’s protocol for SARS-CoV-2 genome sequencing by replacing some primers for the multiplex PCR ^17^. The PCR products in pool 1 and 2 from the same clinical sample were pooled, purified, and subjected to Illumina library construction using a QIAseq FX DNA Library Kit (QIAGEN, Hilden Germany). NextSeq 500 platform (Illumina, San Diego, CA) was used for sequencing the indexed libraries. The next-generation sequencing (NGS) reads were mapped to the SARS-CoV-2 Wuhan-Hu-1 reference genome sequence (29.9 kb ss-RNA; GenBank ID: MN908947), resulting in the specimen-specific SARS-CoV-2 genome sequence by fully mapping on the reference. These mapped reads of SARS-CoV-2 sequences were assembled using A5-miseq v.20140604 ^18^ to determine the full genome sequence (Supplementary Table S2). The single nucleotide variations (SNV) sites and marked heterogeneity were extracted by the read-mapping at ≥10× depth and from 99 - 29796 nt region of Wuhan-Hu-1 genome sequence.

### Comparative genome sequence and SNV analyses

The nearly full-length genome sequence (≥ 29 kb) of SARS-CoV-2 was retrieved from the GISAID EpiCoV database in 28 April 2020, followed by multiple alignment using MAFFT v7.222 ^19^. The poorly aligned regions in 5’ and 3’ end were trimmed; we determined that the core regions were from 99 to 29796 nt position against Wuhan-Hu-1 genome sequences (GISAID ID, EPI_ISL_402125; GenBank ID, MN908947.3). Gap-containing sequences in the core region were excluded; sequences of 2,887 isolates in GISAID database were eventually used in subsequent analyses (isolate collection by 16 April 2020; submitted to GISAID by 27 April 2020; Supplementary Table S3). The genome sequences were aligned using MAFFT software together with sequences retrieved from databases, followed by extraction of SNV sites. The network analysis was performed using TCS 1.21 ^20^.

### Network graph visualization

We had applied various visualization software for network graph visualization such as Network, PopART, TCS, and Cytoscape. Nevertheless, all of the currently available resources have not been suitable for the spatiotemporal dissection of SARS-CoV-2 genomic epidemiology and substantial on-site cluster eradication. Thus, we developed Haplotype Explorer software to visualise the spatiotemporal pattern of SARS-CoV-2 infections based on network graphs (https://github.com/TKSjp/HaplotypeExplorer). Haplotype Explorer allows for the analysis of network graphs spatiotemporally by searching and filtering nodes interactively according to meta-data (e.g., collection date, location, and accession number). Over 100 network graph images were captured daily from the initial detection of SARS-CoV-2 (Wuhan-Hu-1; 31 December 2019) to 16 April, the day when Japan declared a national emergency. Interactive network file (html file; Supplementary dataset 1) and the movie (mp4 file; Supplementary dataset 2) are available.

### Comparison between outdoor activity and number of cases in Japan, the USA, and the UK

We explored the relationships between outdoor activity, daily COVID-19 cases, and local stay-at-home/shelter-in-place order/self-restraint campaigns in Japan, the USA (for New York city and San Francisco Bay area), and the UK. The data shows population mobility normalised to the value of 13 January 2020, which were retrieved from the COVID-19 mobility report provided by Apple (the data is available for a limited time only)^21^. The values for walking were plotted for the three mentioned countries. Subsequently, we plotted the mobility graph with daily and total COVID-19 cases, which were provided by the National Institute of Infectious Diseases (currently the only Japanese data available) ^22^, Coronavirus Data in the United States (provided by New York Times) ^23^, and Coronavirus Source Data ^24^.

## Results and Discussion

The nearly full-length genome sequences (≥ 29 kb, 2,887 entries) of SARS-CoV-2 were retrieved from the GISAID EpiCoV database (collection by April 16, 2020; submitted by April 27, 2020) ^5^. We determined the full genome sequences using 435 clinical specimens in Japan collected until April 6, followed by performing a maximum-likelihood (ML) phylogenetic analysis using genome-wide SNVs to trace potential infection routes (Fig. 1). After the identification of the first COVID-19 case in Japan on 15 January 2020, multiple local COVID-19 clusters were observed. The genome sequences of Japanese isolates were assigned based on phylogeny to China isolates from late February, but nationwide dissemination seemed to already be present based on the ML phylogeny (Fig. 1). At least 4 or 5 main clusters are observed on the ML phylogeny, but this was not always adequately explained by the field epidemiological studies performed by local public health centres about the patient’s information such as nationality, being a Wuhan returnee, and travel history.

**Fig 1.**
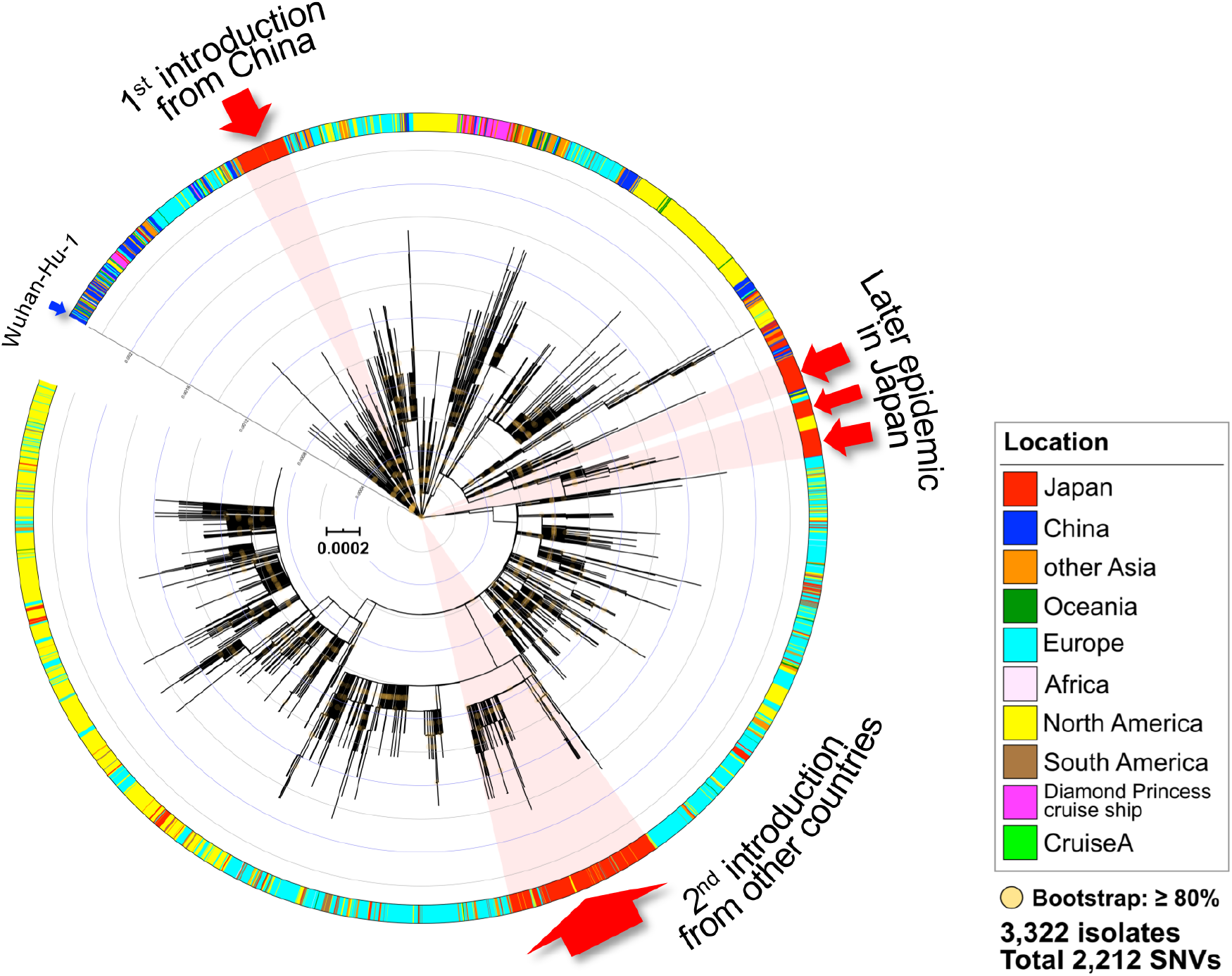
A maximum-likelihood phylogenetic analysis using SARS-CoV-2 genome sequences. In total, 2,212 SNVs were detected in 3,322 isolates, including Japanese isolates (n = 435). The maximum-likelihood phylogenetic tree was constructed using iqtree version 2.0.5 and Wuhan-Hu-1 (GISAID ID: EPI_ISL_402125) as an outgroup reference that is located on the centre point of the radial tree for tree rooting. The geographic and sample information are described in the colour schemes on the outer slot of the phylogenetic tree. The cluster of remarkable outbreaks in Japan are highlighted in bright red.

Although phylogenetic trees are widely used for summarizing genealogies, one should avoid a naїve interpretation of those results since the slow rate of SARS-CoV-2 evolution and sampling bias of genomes could often lead to spurious conclusions ^25^. In order to decode the genealogies of the whole SARS-CoV-2 genome, we performed a haplotype network analysis describing ancestral relationships among the aforementioned genomic data sets and Wuhan-Hu-1 (GISAID ID, EPI_ISL_402125; GenBank ID, MN908947.3) as a potential most recent common ancestral (Fig 2).

**Fig 2.**
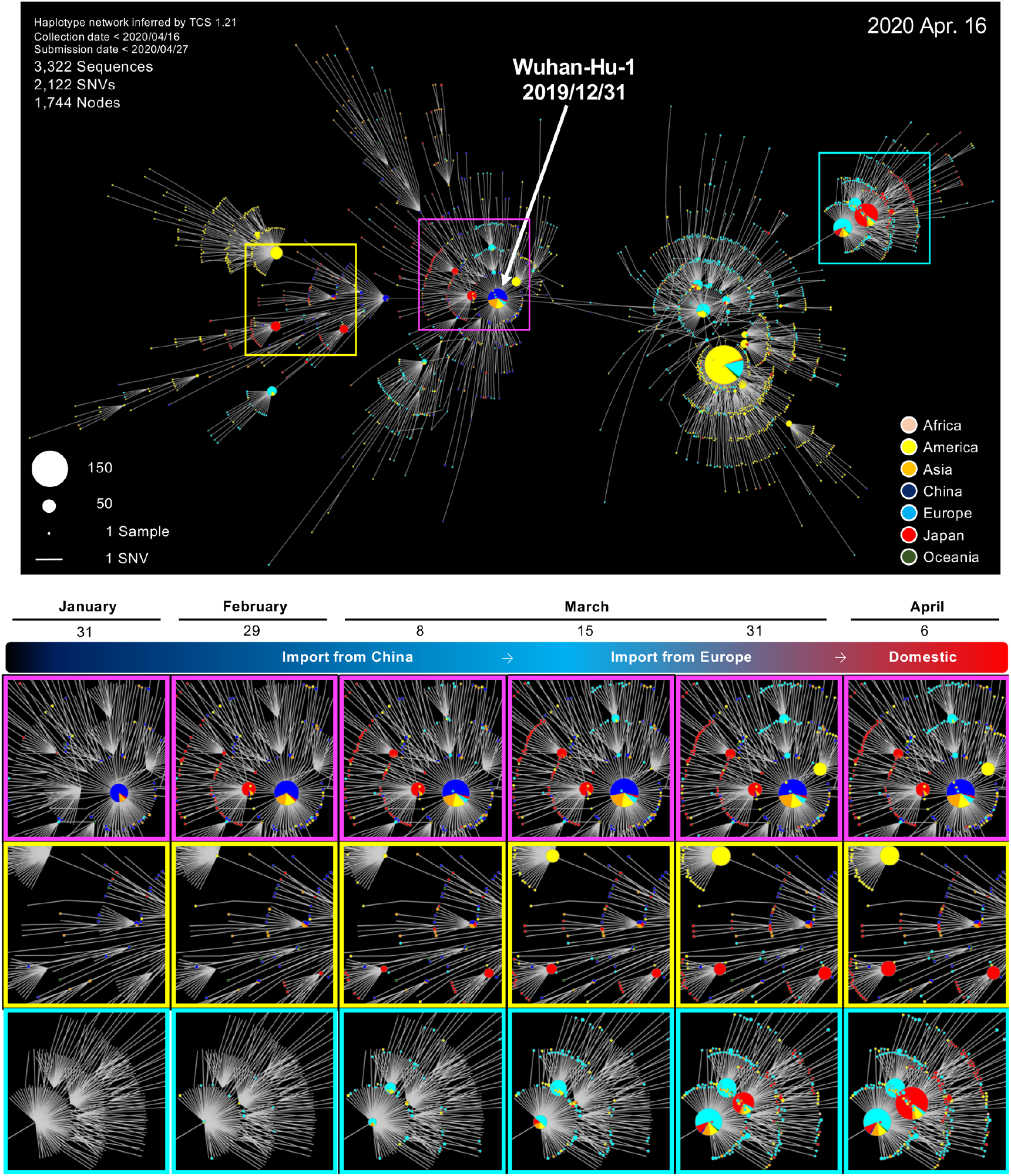
Haplotype network analysis using genome-wide single-nucleotide variations of worldwide SARS-CoV-2 isolates. A) Whole-genome sequences of SARS-CoV-2 isolates in Japan (n = 435) were compared to all GISAID-available SARS-CoV-2 genomes (n = 2,887, updated on 16 April 2020) using TCS network analysis. SARS-CoV-2 disseminating from Wuhan City, China, at the end of December 2019 (one of the potential origins of Wuhan-Hu-1) is plotted at the centre of the haplotype network. B) Three plots of time-series cumulative COVID-19 cases were highlighted in each enclosed square to visualise the increasing incident COVID-19 cases.

Some of the primary clusters identified through January and February in Japan (2 red closed circles and magenta frames in Fig. 2) directly descended from the haplotype which was mostly seen in Wuhan-Hu-1-related isolates from China. Three other distinct clusters (additional 2 red closed circles and yellow frames in Fig. 2) were observed after the first introduction of Chinese isolates. Those four clusters of COVID-19 infections were assumed to be directly originated from the primary wave which occurred in China. Those clusters seemed to have been almost contained through the implementation of active surveillance in Japan until mid-March.

In contrast, the number of COVID-19 cases increased rapidly across Europe and North America during early March (right half of Fig. 2), indicating a pandemic was occurring. Concurrently, many sporadic COVID-19 cases were detected in Japan from the end of March through early April. The haplotype network analysis demonstrated that an additional large cluster (red closed circles and cyan frames in Fig 2) closely related to the outbreak in Europe (originating in China or other countries) was predominantly observed in Japan during early April, suggesting that they could have caused the increased spread within Japan.

Analysis of whole genome sequences provides information that is useful for tracing the spread of infection using genome-wide SNVs among clusters. To elucidate which Japan political decisions (Fig. 3A, and Supplementary Table S4) and activities were effectively involved in the introduction of specific SARS-CoV-2 lineages, we characterised daily COVID-19 case reports and a mobility index of people inferred from iPhone tracking (Fig. 3B) in Japan, USA, and UK before and after the self-restraint campaign.

**Fig 3.**
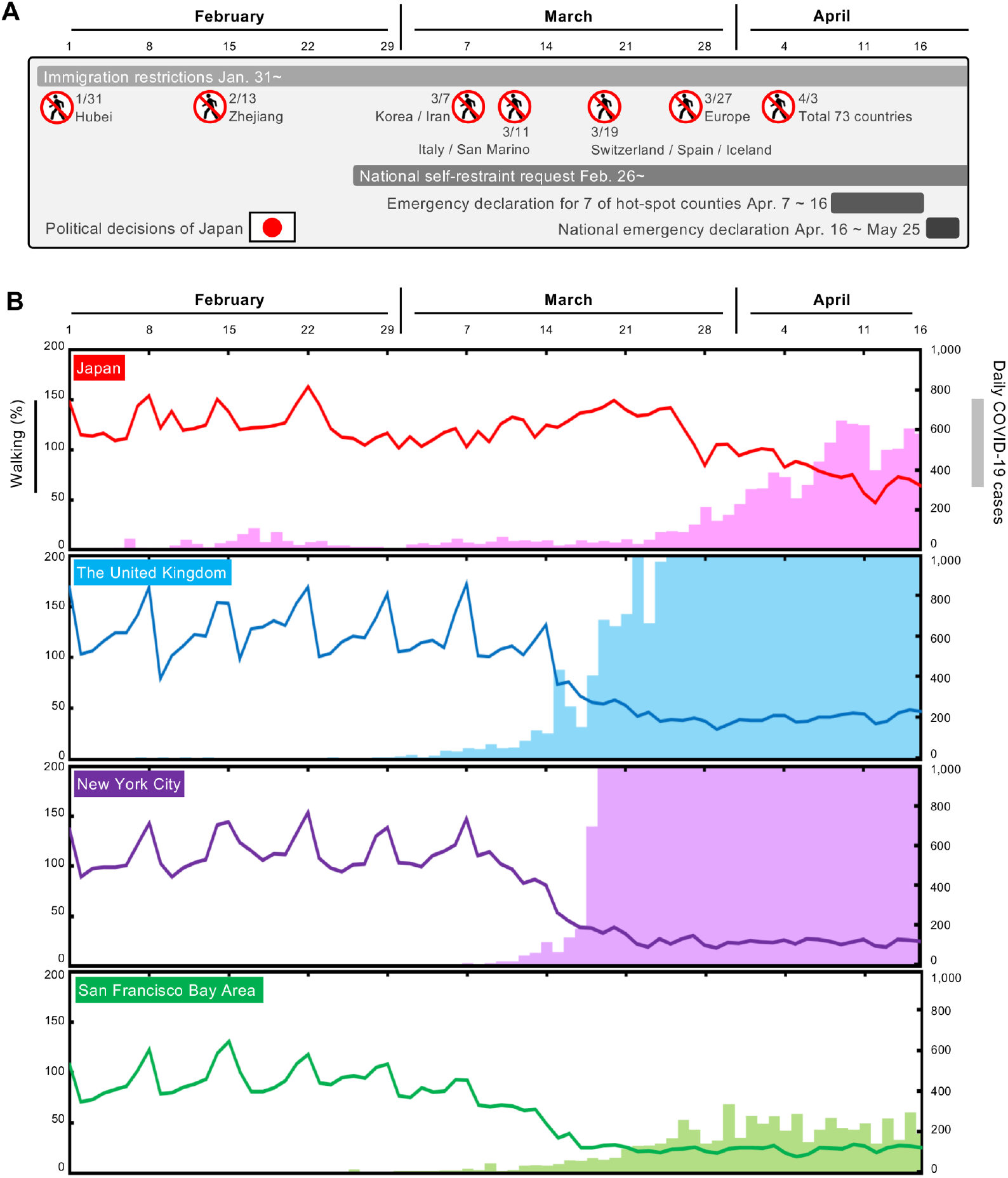
Mobility index (walking) of people and daily COVID-19 cases in Japan, the United Kingdom, New York city, and San Francisco Bay area. A) Timeline of political decisions for the COVID-19 quarantine and national actions in Japan (see also Supplementary Table S4). B) Mobility index of people and daily COVID-19 cases in Japan, the UK, and the USA (New York city and San Francisco Bay Area). Solid lines indicate mobility index of people inferred from map application usages given by Apple. Bar plot indicates daily COVID-19 cases. Notably, the data suggests that people in Japan and San Francisco Bay Area collaborated with “stay-at-home” measures from the end of February, which might have significantly reduced the expansion of SARS-CoV-2 infections after late March 2020.

During the primary wave from China, international airports investigated potential patients with COVID-19 using the keywords “Wuhan, Hubei, Zhejiang, and China” in early February (Fig. 3A), and local public health centres were able to identify patients likely to be infected with COVID-19 and their close contacts within location-specific clusters by conducting active epidemiological surveillance. After the end of February, a national self-restraint was requested in Japan, leading to significant reductions in activity observed after the self-restraint was implemented. Although the situation in Japan had begun to improve around mid-March, a large number of new patients with COVID-19 were diagnosed, many of whom with unclear infection routes. Tracing the infection routes was difficult because some Japanese cases had no recent history of travel to China or any other country outside Japan.

During the national holidays from 20-22 March, a part of the population left their homes to visit the cherry blossoms, which possibly increased mobility (Fig. 3B). It was speculated that such a partial increase in activity at that time might have allowed for a resurgence of the remaining lineage from the first introduction which had previously circulated. The SARS-CoV-2 haplotype network analysis, however, suggested that the second wave of COVID-19 cases in late March had a distinct origin from the first wave lineage. The lineage of the second wave could have been imported by the returnees or travellers from Europe, North America, or other countries ^26^ (see the image in cyan frame in Fig. 2), but not by the remaining Wuhan lineage. Considering the increasing COVID-19 cases in Europe and the detection of imported cases in Japan, the Japanese government decided to stop immigration from Europe in 27 March (Fig. 3A).

On the other hand, increased mobility in the USA and UK continued through early March, possibly leading to a sharp increase in COVID-19 cases from mid-March (Fig. 3B). Intriguingly, activity in the San Francisco Bay area slowed down earlier than in New York; the additional 3 days taken by New York to implement stay-at-home/shelter-in-place orders compared to San Francisco may explain the mitigated COVID-19 spread in San Francisco.

## Conclusions

This genome surveillance study suggested that at least two distinct SARS-CoV-2 introductions from China and other subsequent countries, including some in Europe, were detected. To mitigate the next wave of COVID-19 in Japan, it is necessary to formulate a more efficient containment strategy using real-time genome surveillance supporting epidemiological field investigations and highlight potential infection linkages.

## Data Availability

The new sequences have been deposited in Global Initiative on Sharing All Influenza Data (GISAID) with accession IDs EPI_ISL_479792 to EPI_ISL_480227 (Table S2).

https://www.gisaid.org/epiflu-applications/next-hcov-19-app/

## Acknowledgments

We sincerely thank the COVID-19 Genomic Surveillance Network in Japan (Supplementary Table S1) for the collection and transportation of clinical specimens. We are particularly grateful to the staff of the National Institute of Infectious Diseases, Field Epidemiology Training Program (FETP) team; the Ministry of Health, Labor and Welfare; and local governments for their assistance with administrative matters, field investigations, data collection, and laboratory testing.

We would like to thank all researchers who have kindly deposited and shared genomic data on GISAID. A table with genome sequence acknowledgments can be found in Supplementary Table S3.

## Author Contributions

TS1, KI, WT, and MK designed and organised the genome study.

AO, KG, HT, HE, KS, MT, SS, RN, SH, MT, and COVID-19 Genomic Surveillance Network in Japan (Table S1) performed the laboratory detection.

RT and MH performed the genome sequencing, and TS1, KI, TKS and KY performed the genome analysis.

TS2, TM, TS3, HK, YY, TY, and MS contributed to the field epidemiological study. MK wrote the manuscript.

## Declaration of interests

The authors declare that the research was conducted in the absence of any commercial or financial relationships that could be construed as a potential conflict of interest.

**Supplementary Table S1**

Collaboration with local public health institutes in COVID-19 Genomic Surveillance Network in Japan.

**Supplementary Table S2**

Summary of NGS reads and in silico data analysis.

**Supplementary Table S3**

All GISAID-available SARS-CoV-2 genomes (n = 2,887).

**Supplementary Table S4**

Japanese immigration restriction programs.

**Supplementary dataset 1**

Interactive genomic network file (html format).

**Supplementary dataset 2**

Genomic network movie file (mp4 format).

